# Differences in coagulation responses to vascular injury between uninterrupted dabigatran and apixaban - a clinical prospective randomized study

**DOI:** 10.1101/2023.12.18.23300179

**Authors:** Yasuhiro Ikami, Daisuke Izumi, Shinya Fujiki, Hirotaka Sugiura, Sou Otsuki, Naomasa Suzuki, Yuta Sakaguchi, Takahiro Hakamata, Yuki Hasegawa, Nobue Yagihara, Kenichi Iijima, Takahiro Tanaka, Masahiro Ishizawa, Masaomi Chinushi, Tohru Minamino, Takayuki Inomata

## Abstract

**Background:** The coagulation response during vascular injury with uninterrupted administration of direct oral anticoagulants (DOACs) has not been elucidated. Our aim was to evaluate differences in coagulation responses after vascular injury between uninterrupted direct thrombin inhibitor and direct factor Xa inhibitor recipients.

**Methods:** Patients scheduled for catheter ablation for atrial fibrillation were randomly assigned to receive dabigatran or apixaban in this prospective, randomized, comparative, parallel-group study. Venous blood was collected three times: 180 minutes after taking the anticoagulant on the day before the procedure, before vascular punctures of the ablation procedure, and 10–15 minutes after the start of vascular punctures.

**Results:** Forty-two patients were enrolled. The prothrombin fragment 1+2 (F1+2) level, the primary endpoint, was much larger after vascular puncture in the uninterrupted dabigatran recipients (median: 83 pmol/L; interquartile range: 56–133 pmol/L) than in the uninterrupted apixaban recipients (median: 1 pmol/L; interquartile range: −3–19 pmol/L; P < 0.001). Antithrombin levels decreased after vascular puncture in dabigatran recipients, and both protein C and antithrombin levels decreased after vascular puncture in apixaban recipients.

**Conclusions:** Unlike uninterrupted apixaban, uninterrupted dabigatran does not inhibit thrombin generation in response to vascular injury.

**Clinical perspective:** *What is new?:*

- To the best of our knowledge, this is the first randomized clinical comparison of the effects of direct thrombin and factor Xa inhibitors on the physiological coagulation and anticoagulation system after vascular injury while direct oral anticoagulants (DOACs) serum levels were at the peak phase.
- Unlike uninterrupted apixaban, uninterrupted dabigatran does not inhibit thrombin generation in response to vascular injury.
- This study shows that physiological anticoagulation factors are consumed during vascular injury in patients receiving DOACs.

*What are the clinical implications?:*

- The difference in the thrombin generation response during vascular injury between uninterrupted dabigatran and apixaban may be one of the reasons for different clinical outcomes of thrombotic and hemorrhagic complications.
- DOACs may inhibit an excessive coagulation response by retaining physiological anticoagulation factors.

## Introduction

Atrial fibrillation (AF) has major clinical implications for patients’ quality of life, morbidity with ischemic stroke and heart failure, and mortality (**1–6**). Anticoagulation therapy is recommended for patients with risk factors for thromboembolism or during the perioperative period of catheter ablation for AF (**7,8**). Direct oral anticoagulants (DOACs), which carry a lower risk of bleeding and interact less with diet than other anticoagulants, are commonly used for anticoagulation therapy (**9–11**). Uninterrupted administration of DOACs is becoming more common during the perioperative period of catheter ablation for AF because recent randomized clinical studies of such treatment showed that the risk of major bleeding was lower than or similar to that with uninterrupted warfarin treatment (**12–15**). In addition, in these randomized controlled trials, in which DOACs and warfarin were administered during the perioperative period of AF ablation, the frequency of hemorrhagic complications among patients who received dabigatran was conspicuously low (**13**). However, the reason for the low risk of major bleeding complications when catheter ablation is performed during the peak phase of dabigatran is unclear (**13**). Therefore, this multicenter, randomized study evaluated differences in coagulation responses, including thrombin production after vascular injury, between direct thrombin inhibitor and direct factor Xa inhibitor recipients.

## Methods

### Study Design

This prospective, open-label, randomized, comparative, parallel-group study was conducted at two centers, Niigata University Medical and Dental Hospital and Niigata Medical Center, from August 2019 to March 2021. It was performed in accordance with the guiding principles of the Declaration of Helsinki and in compliance with the Clinical Trial Act, a Japanese law designed to ensure that researchers monitor and adhere to the study criteria. The Niigata University Clinical Research Central Review Board approved the study protocol (number SP18014), which was included in the Japan Registry for Clinical Trials (ID jRCT1031190030). Informed consent was obtained from all patients in accordance with these regulations. The institutional review board comprehensively conducted an ethical review of all participating facilities.

### Participants

Eligible patients were 40–80 years of age, scheduled for catheter ablation for paroxysmal or persistent AF according to the JCS/JHRS 2019 Guideline on Non-Pharmacotherapy of Cardiac Arrhythmias (**7**), were eligible for treatment with dabigatran (a direct thrombin inhibitor) or apixaban (a direct factor Xa inhibitor) according to the prescribing guidelines in Japan (**9**), and provided informed consent to participate. The main exclusion criteria were (1) a history of intracranial hemorrhage or bleeding in other important organs; (2) the presence of mechanical heart valves or hemodynamically significant mitral valve stenosis; (3) moderately severe or worse hepatic dysfunction (Child-Pugh class B or C); (4) renal dysfunction characterized by a Cockroft–Gault estimated creatinine clearance <50 mL/min; (5) conditions with a high risk of bleeding (e.g., uncontrolled severe hypertension, active malignant tumors, congenital or acquired bleeding disorders, active ulcerative gastrointestinal disorders, treatment with two or more antiplatelet agents); and (6) a New York Heart Association class III or worse heart failure, left ventricular ejection fraction of 35% or less according to echocardiography, or a history of hospitalization for heart failure within 1 year before the study.

### Interventions

#### Patient assignment

Participants were randomly assigned in a 1:1 ratio to receive dabigatran or apixaban, and the random allocation sequence was computer generated by a research physician. Dynamic allocation with a minimization method was used to assign patients. The adjusting factors were age, gender, and CHADS_2_ score (calculated as 1 point each for a history of hypertension, diabetes, recent heart failure, and age ≥75 years and 2 points for a history of a stroke or transient ischemic attack) (**16**). A comprehensive medical history review, physical examination, blood tests, and transthoracic echocardiography were performed after obtaining consent from the participants.

#### The dose of the designated anticoagulants

If an anticoagulant drug different from the designated drug had been used before the study, it was discontinued before drug assignment. After the drug assignment, we administered the designated DOACs, and administration continued until after catheter ablation for AF. Dabigatran was administered at a dose of 150 mg twice daily or, if dosage adjustment was required (e.g., for patients aged ≥75 years, those taking P-glycoprotein inhibitors, or those with a history of gastrointestinal bleeding), 110 mg twice daily. Apixaban was administered at a dose of 5 mg twice daily or, if dose adjustment was required (≥2 of the following criteria: age of ≥80 years, body weight ≤60 kg, or serum creatinine ≥1.5 mg/dL), 2.5 mg twice daily. The designated DOACs were administered at least 1 week before catheter ablation for AF.

#### Examinations during hospitalization, including blood sampling before and after vascular puncture

Participants were admitted to the hospital two days before the ablation procedure. Baseline measurements, physical examinations, blood tests, electrocardiography, and transthoracic echocardiography were performed at admission. Venous blood was collected three times to investigate the effect of vascular injury on the coagulation system: on the day before the procedure, 180 minutes after the DOAC was administered (i.e., when serum levels of the DOAC are at the peak phase) (**17**); before vascular puncture for the ablation procedure; and 10–15 minutes after the start of vascular puncture, which was performed via a venous sheath introducer after all sheath introducers were inserted and before intravenous administration of unfractionated heparin. We planned not to use data from patients who required more than 15 minutes to insert the sheath introducers. The final DOAC dose was administered 1-2 hours before the ablation procedure (Figure 1).

**Figure 1.**
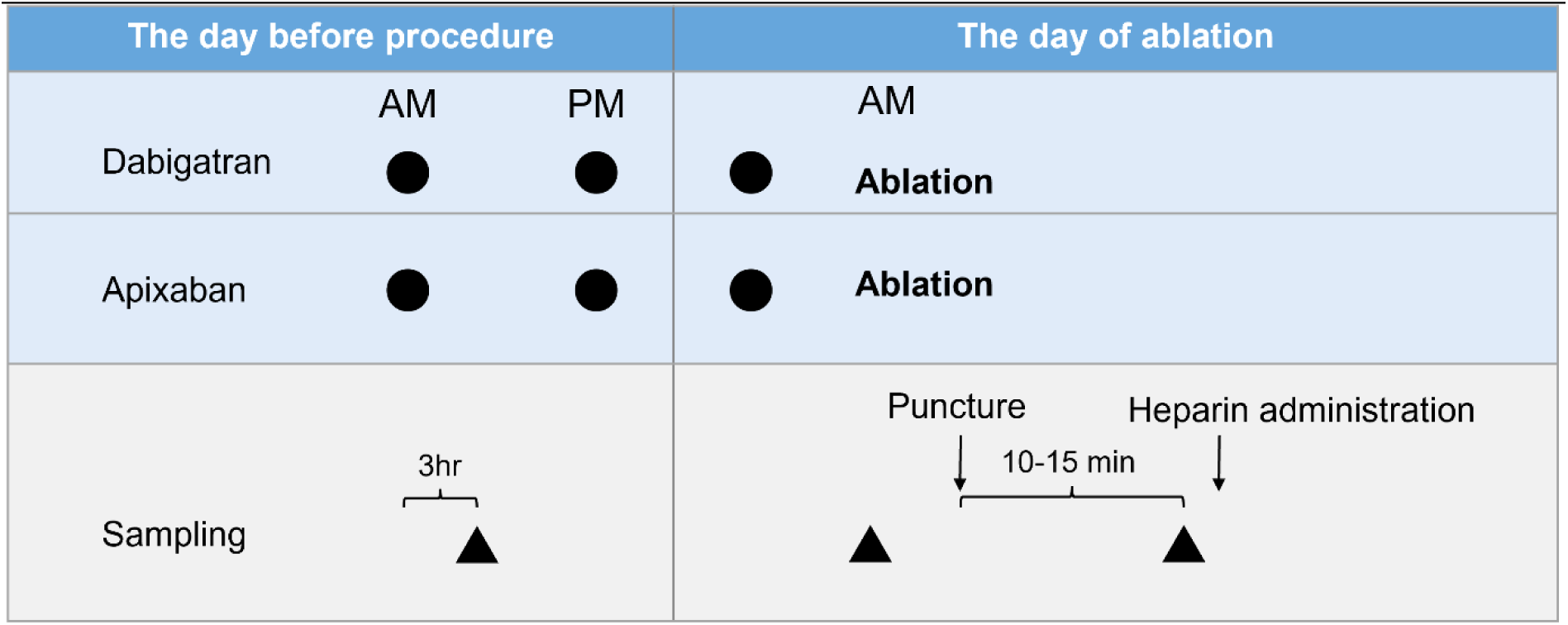
Study protocol. Venous blood was collected thrice to investigate the effect of vascular injury on the coagulation system in uninterrupted DOAC recipients: 180 minutes after the anticoagulant was administered on the day before the procedure, before the vascular punctures of the ablation procedure, and 10–15 minutes after the initiation of the vascular punctures.

The vascular puncture was performed according to the modified Seldinger method. Conventional anatomical landmarks, palpation, and real-time two-dimensional vascular echocardiography were used to locate the internal jugular veins, femoral arteries, and femoral veins (**18,19**). In all patients undergoing AF ablation, four or five vascular sheath introducers were inserted: one or two 8 Fr (SL0; St. Jude Medical Japan Co. Ltd., Tokyo, Japan) and one 8.5 Fr (SR0; St. Jude Medical Japan Co. Ltd.) into the femoral vein; one 7.2 Fr (Medikit Supersheath; Medikit Co. Ltd., Tokyo, Japan) into the internal jugular vein; and one 3 Fr (XEMEX, Zeon Medical, Tokyo, Japan) into the femoral artery.

We evaluated the prothrombin fragment 1+2 (F1+2) levels as a marker of thrombin generation, soluble fibrin monomer complex (SFMC) as a marker of coagulation activation, and D-dimer as a marker of coagulation. Similarly, we evaluated protein C, protein S, and antithrombin levels as physiological coagulation inhibitors. Sample measurements were made at an external institution unrelated to this study. We used an ELISA (Enzygnost F1+2 [Monoclonal]; Siemens Healthcare Diagnostic Corporation, Tokyo, Japan) to determine the F1+2 level, a hemagglutination assay (FM test; Fujirebio Inc., Tokyo, Japan) or latex photometric immunoassay (Iatro SF II; LSI Medience Co., Tokyo, Japan) to quantify SFMC level, and functional assays (HemosIL ProClot and HemosIL PS-clot, respectively, I.L. Japan Co., Ltd., Tokyo, Japan) to quantify protein C and S levels. We used the chromogenic substrate method (Testzym S ATIII, Sekisui Medical Co., Ltd., Tokyo, Japan) to measure antithrombin activity and a latex agglutination assay to measure D-dimer levels (LATECLE D-dimer, KAINOS Laboratories, Inc., Tokyo, Japan).

### Endpoints

The primary endpoint of this study was the change in F1+2 levels before and after vascular puncture for both DOACs. Secondary endpoints were F1+2, protein C, protein S, and antithrombin levels and the changes in the latter three levels between vascular punctures for both DOACs. The exploratory endpoints were all D-dimer levels and the changes in D-dimer and SFMC levels between vascular punctures for both DOACs. All adverse events during this study were recorded, analyzed, and classified as serious or nonserious. The events were reported to the Data and Safety Monitoring Committee and evaluated for recommendations to continue the study.

### Statistical Analysis

Based on previous findings (20), we hypothesized that the increase in F1+2 levels before and after vascular puncture would be approximately 2.5 times higher for dabigatran than for apixaban. Because we used 80% power and 0.05 bilateral alpha levels, the study had to include at least 36 patients to reach statistical significance. Given the potential number of dropouts, we set the target number of participants at 40.

The safety analysis set included all enrolled patients. The full analysis set included all enrolled patients except those with critical protocol violations (e.g., no consent or major procedural violations). The per protocol set included all enrolled patients from the full analysis set without major protocol deviations.

The study endpoints were analyzed for the full analysis set. Continuous variables were calculated as means and standard deviations for normally distributed data and as medians and interquartile ranges (IQRs) for nonnormally distributed data. Categorical variables were calculated as numbers (percentages). Patient characteristics were compared with chi-square tests (for five or more events) and Fisher’s exact test (for fewer than five events) for categorical variables, *t*-tests for normally distributed continuous variables, and the Wilcoxon rank-sum test for continuous variables with a skewed distribution. All *P*-values were two-sided, and values <0.05 were statistically significant. We used SPSS Statistics 27 (IBM Corporation, Armonk, NY, USA) to perform all statistical analyses.

## Results

### Patient Characteristics

Of the 42 patients randomly assigned to receive dabigatran or apixaban, 1 patient assigned to receive dabigatran withdrew from the study before drug administration. The trial was terminated because the target number of enrolled patients was reached. Persistent AF was found in nine patients (45%) who received dabigatran and seven patients (33%) who received apixaban. Thirty-seven patients (17 receiving dabigatran and 20 receiving apixaban) completed the study. However, one patient receiving dabigatran was excluded from the per protocol set because of a protocol violation (a mistake in the blood sampling procedure). There were no changes to trial outcomes after the trial commenced. The demographic and clinical characteristics of the participants of both groups were similar (Table 1).

**Table 1.** Baseline characteristics.

Baseline characteristics
|  | **Dabigatran group (n=20)** | **Apixaban group (n=21)** | **P value** |
| --- | --- | --- | --- |
| **Female, n (%)** | 4 (20) | 4 (19) | 1 |
| **Age, years** | 61.3 ± 8.5 | 63.8 ± 7.8 | 0.34 |
| **Type of atrial fibrillation, n (%)** |  |  | 0.44 |
| **Paroxysmal** | 11 (55) | 14 (67) |  |
| **Persistent** | 9 (45) | 7 (33) |  |
| **Chronic heart failure, n (%)** | 0 | 0 | 1 |
| **Hypertension, n (%)** | 11 (55) | 14 (67) | 0.44 |
| **Diabetes, n (%)** | 2 (10) | 3 (14) | 1 |
| **History of stroke or transit**<br>**ischemic attack, n (%)** | 2 (10) | 1 (5) | 0.61 |
| **CHADS2 score** | 0.85 ± 0.81 | 0.95 ± 0.67 | 0.66 |
| **Low-dose anticoagulants, n (%)** | 2 (10) | 0 | 0.23 |
| **Body weight, kg** | 72.0 ± 13.4 | 73.4 ± 13.1 | 0.74 |
| **Systolic blood pressure, mmHg** | 126 ± 16 | 123 ± 14 | 0.57 |
| **Diastolic blood pressure, mmHg** | 80 ± 12 | 77 ± 11 | 0.50 |
| **Heart rate, bpm** | 76 ± 12 | 71 ± 11 | 0.18 |
| **Body temperature, °C** | 36.2 ± 0.4 | 36.3 ± 0.6 | 0.53 |
| **Left ventricular ejection fraction, %** | $63.7 \pm 6.7$ | $63.2 \pm 6.7$ | 0.80 |
| **Aspartate aminotransferase, U/L** | $24 \pm 7$ | $28 \pm 9$ | 0.14 |
| **Alanine aminotransferase, U/L** | $25 \pm 9$ | $31 \pm 13$ | 0.10 |
| **Lactate dehydrogenase, U/L** | $191 \pm 23$ | $194 \pm 25$ | 0.72 |
| **Creatinine, mg/dL** | $0.83 \pm 0.13$ | $0.83 \pm 0.15$ | 0.90 |
| **Estimated creatinine clearance, mL/min** | $94.1 \pm 22.2$ | $92.7 \pm 22.1$ | 0.85 |
| **Hemoglobin A1c, %** | $5.8 \pm 0.4$ | $5.8 \pm 0.5$ | 0.83 |
| **Hemoglobin, g/dL** | $14.7 \pm 1.4$ | $14.8 \pm 1.2$ | 0.81 |

### Vascular Punctures

We found no significant difference between the two groups from the final DOAC administration to the blood samplings (Table 2). In addition, the number of sheath introducers inserted did not differ significantly between the two groups (Table 2).

**Table 2.**
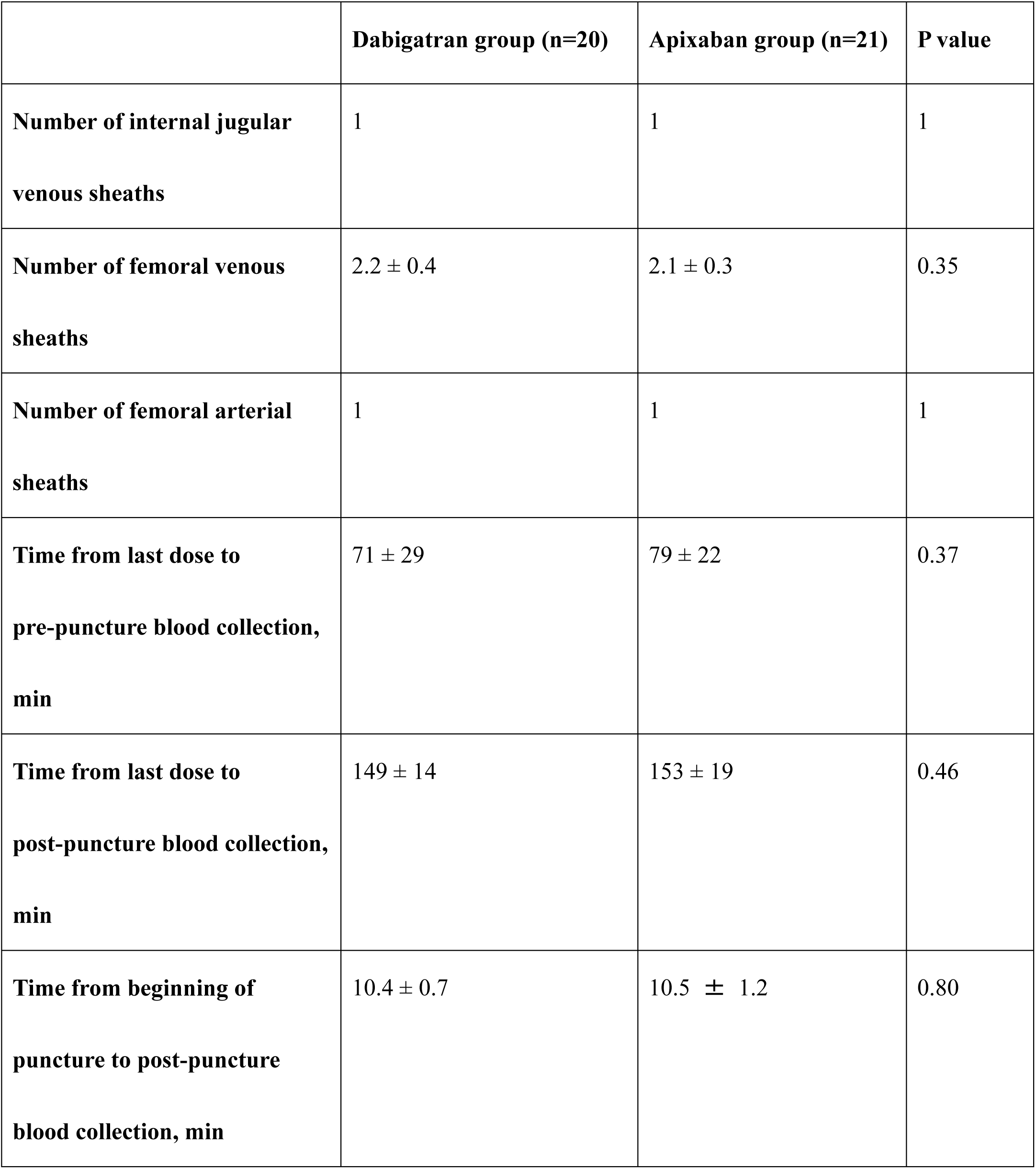
Vascular puncture situation.

### Attrition and Adverse Events

During the study period, five patients (four receiving dabigatran and one receiving apixaban) dropped out (Figure 2). Of those receiving dabigatran, 1 voluntarily withdrew consent, 1 patient dropped out because of upper gastrointestinal symptoms, and 2 suffered unexpected adverse events during ablation procedure (sedative-induced coronary spasm in 1, delay of 15 minutes or more in vascular puncture in the other). Of those receiving apixaban, 1 patient withdrew because the catheter ablation procedure was canceled. Of those receiving dabigatran, 2 suffered adverse events that may have been caused by the study (upper gastrointestinal tract symptoms in 1 and gastric ulcer associated with discontinuation of proton pump inhibitor therapy in 1). Thus, the per protocol set included 36 patients.

**Figure 2.**
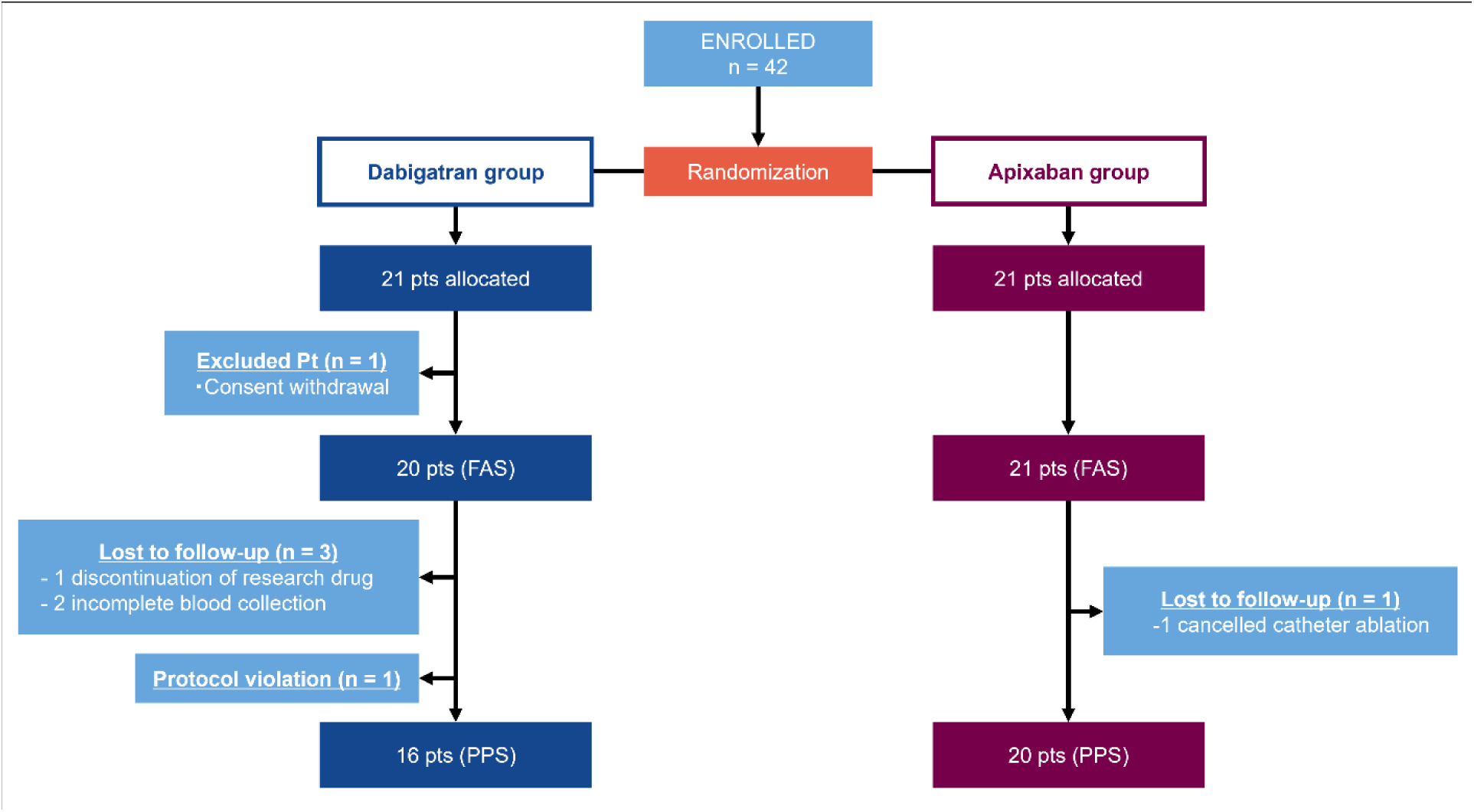
Flowchart of randomized participant assignment and the data analyzed. FAS indicates full analysis set; PPS, per protocol set.

### Primary Endpoint

The change in F1+2 levels after vascular punctures was much larger in participants taking dabigatran (median: 83 pmol/L [IQR: 56 to 133 pmol/L]) than in those taking apixaban (median: 1 pmol/L [IQR: −3 to 19 pmol/L] pmol/L]; *P* < 0.001; Figure 3).

**Figure 3.**
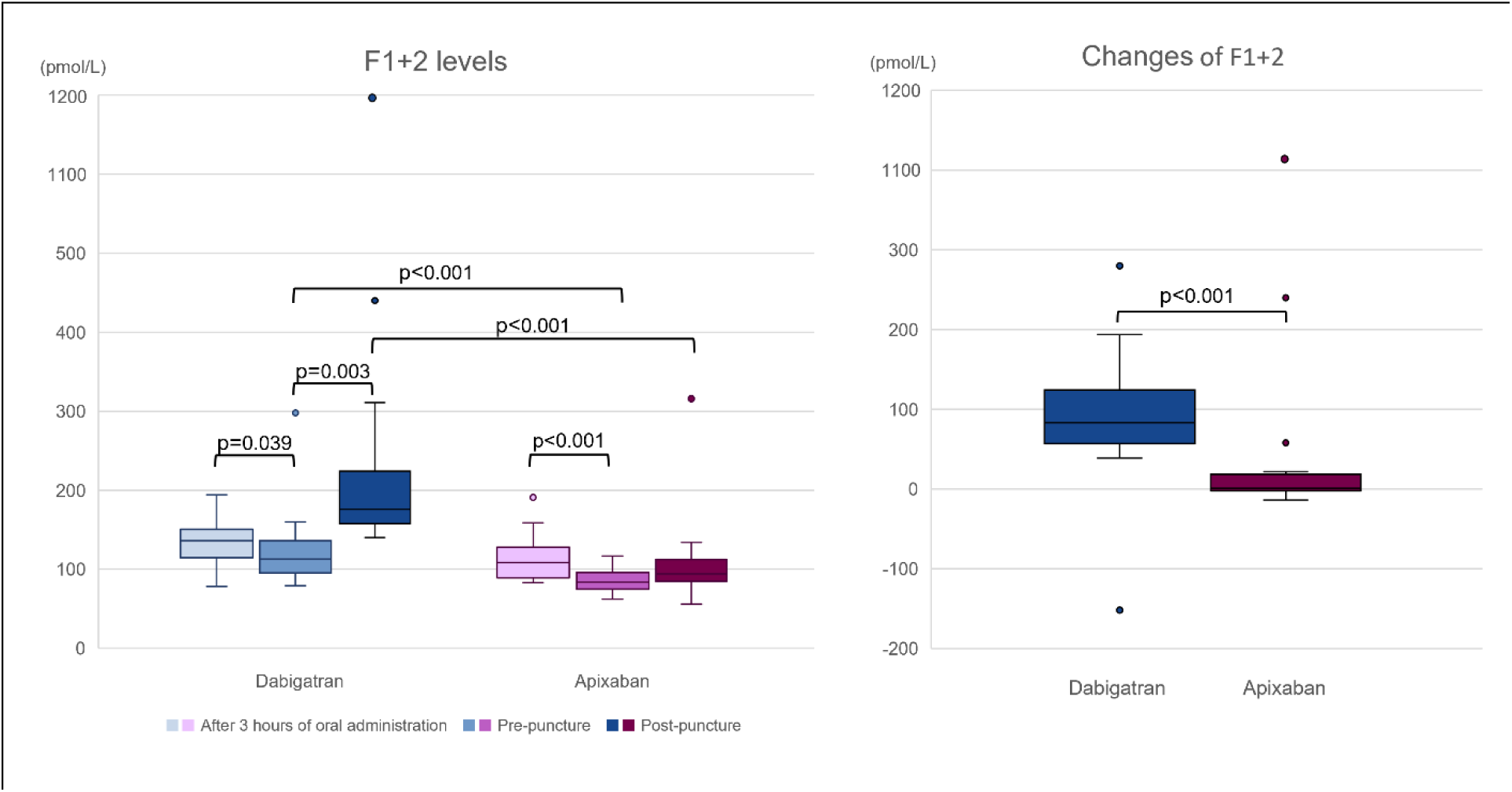
Prothrombin fragment 1+2 (F1+2) levels before and after vascular puncture and the changes in F1+2 levels after vascular punctures. Circles represent measurements outside the 1.5 × interquartile range. F1+2 indicates prothrombin fragment 1+2.

### Secondary Endpoints

F1+2 levels before vascular puncture were slightly higher in the dabigatran recipients (median: 113 pmol/L [IQR: 93 – 141 pmol/L]) than in the apixaban recipients (median: 84 pmol/L [IQR: 72 – 96 pmol/L]; *P* < 0.001). Unlike the apixaban recipients, dabigatran recipients showed a significant increase in F1+2 levels after vascular puncture (Figure 3). Antithrombin levels were significantly higher in the apixaban recipients before and after vascular puncture. Moreover, protein C and S levels were significantly higher in the dabigatran recipients before and after puncture (Figure 4). In both groups, F1+2 levels before vascular puncture were slightly lower than 3 hours after the administration of DOACs on the day before the procedure, when serum DOAC levels are in peak phase; in dabigatran recipients, median levels were 113 pmol/L (IQR: 93 to 141 pmol/L) vs. 136 pmol/L (IQR: 112 to 151 pmol/L), respectively (*P* = 0.039); in apixaban recipients, median levels were 84 pmol/L (IQR: 72 to 96 pmol/L) vs. 109 pmol/L (IQR: 88 – 128 pmol/L), respectively (*P* < 0.001) (Figure 3). In the dabigatran recipients, antithrombin levels decreased after vascular puncture, whereas in apixaban recipients, protein C and antithrombin levels decreased after vascular puncture (Figure 4).

**Figure 4.**
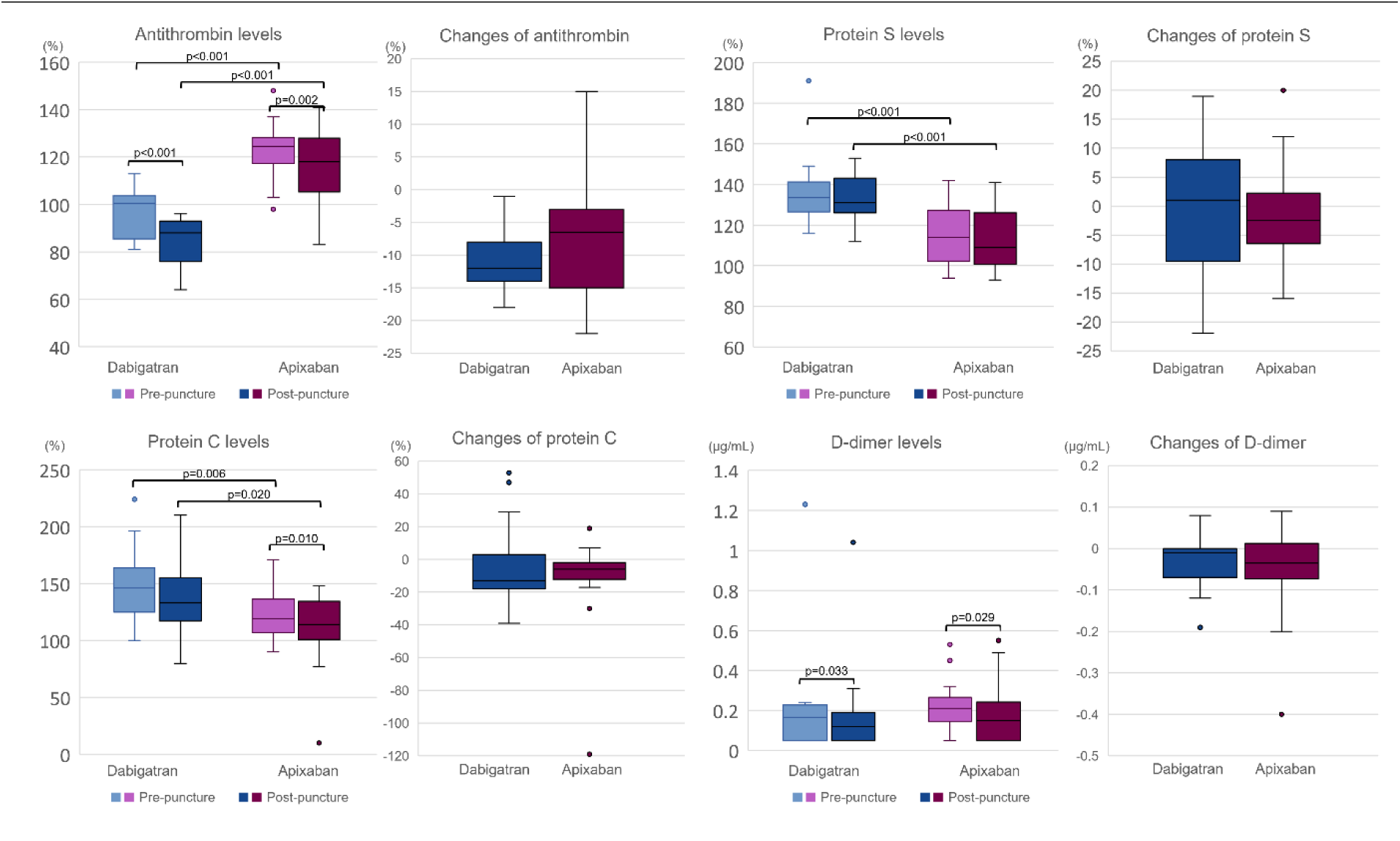
Antithrombin, protein S, protein C, and D-dimer levels before and after vascular punctures and the changes in these levels after vascular punctures. Circles represent measurements outside the 1.5 × interquartile range.

### Exploratory Endpoints

We found no significant difference in pre-puncture D-dimer levels between the dabigatran (median: 0.17 μg/mL [IQR: 0.05 to 0.23 μg/mL]) and apixaban recipients (median: 0.21 μg/mL [IQR: 0.14 to 0.30 μg/mL]; *P* = 0.067) or in post-puncture D-dimer levels between the dabigatran (median: 0.12 μg/mL [IQR: 0.05 to 0.23 μg/mL]) and the apixaban recipients (median: 0.15 μg/mL [IQR: 0.05 to 0.25 μg/mL]; *P* = 0.42). We also found no significant difference in the change in D-dimer levels between the dabigatran (median: −0.01 μg/mL [IQR: −0.08 to 0 μg/mL]) and the apixaban recipients (median: −0.04 μg/mL [IQR: −0.08 to 0.02 μg/mL]; *P* = 0.92; Figure 4). Meanwhile, the SFMC changed positively (*P* = 0.58) after vascular puncture in two dabigatran recipients and one apixaban recipient. SFMC was measured with two different methods, but the difference in the method did not affect the change in SFMC (*P* > 0.999).

## Discussion

Our major findings were that (1) after vascular puncture, F1+2 levels increased more in the uninterrupted dabigatran recipients than in the uninterrupted apixaban recipients, and (2) Some coagulation inhibitors, such as antithrombin and protein C, appeared to be consumed after vessel puncture.

To our knowledge, this is the first randomized clinical comparison of the effects of direct thrombin and factor Xa inhibitors on the physiological coagulation and anticoagulation system after vascular injury while serum levels of DOAC were at the peak phase. Several previous studies of the anticoagulant effect of DOACs have shown that thrombin generation or levels of coagulation activation markers, such as F1+2 and soluble fibrin, are suppressed to some extent not only during the peak phase but also during the trough phase of DOACs (**20–23**). The degree of suppression of F1+2 levels at the peak levels of DOAC in our study was similar to those previously reported (**20–23**). According to our previous report, thrombin production was increased at dabigatran and apixaban trough levels during vascular injury, but the response at DOAC peak levels was unknown (**20**). In this study, we found that the large thrombin production after vascular puncture during the peak phase of dabigatran was similar to the trough phase previously reported (**20**). Meanwhile, we found that an increased response to thrombin generation after vascular puncture was not observed during the peak phase of apixaban (**20**).

Unlike direct Xa inhibitors, direct thrombin inhibitors in the therapeutic range may be less effective in suppressing initial thrombin generation associated with vascular injury (**24**). Moreover, direct factor Xa inhibitors in the therapeutic range may suppress thrombin bursts after the initial thrombin generation (**25**). This finding may be the reason for the difference in thrombin-producing ability after vascular injury observed in this study. Compared with apixaban, dabigatran appeared to inhibit hemorrhagic complications such as intracranial hemorrhage and bleeding complications during catheter procedures (**12–15,26**). In contrast, the risk of thrombotic events such as asymptomatic cerebral infarction during ablation may be greater in patients taking dabigatran (**15,23,27**). The difference in the thrombin generation response between both drugs observed in our study may be one of the reasons for the different clinical outcomes of thrombotic and hemorrhagic complications mentioned above.

Our results suggest that dabigatran increased the protein C and S levels, whereas apixaban increased antithrombin levels. DOACs may prolong clotting time, and protein C, protein S, and antithrombin activity may be overestimated (**28,29**). Meanwhile, antithrombin or protein C levels decreased after vascular puncture in both groups in this study. Although there is an interaction of DOACs on the measured values, before-and-after comparisons are possible. Therefore, decreased physiological anticoagulant factors during vascular puncture may indicate negative feedback for a hypercoagulable state in the DOAC recipients.

Our study had some limitations. The sample size was relatively small, and the study was underpowered to determine the difference in the increasing reaction of the SFMC. Blood samples obtained before vascular puncture might not accurately reflect the peak levels of DOACs because the elapsed time from the last administration of DOACs was approximately 70–80 minutes (**30,31**). However, F1+2 levels 3 hours after the administration of DOACs at admission, when DOAC levels peaked, were equivalent to or higher than F1+2 levels before vascular puncture. Therefore, the comparison of each measurement before and after vascular puncture in this study is considered an evaluation, while DOAC levels are fairly high.

## Conclusions

In contrast to uninterrupted apixaban, uninterrupted dabigatran did not inhibit thrombin generation in response to vascular injury.

## Data Availability

For privacy purposes, the data that support the findings of this study are available on request from the corresponding author.

## Non-standard Abbreviations and Acronyms

AF: Atrial fibrillation
DOAC: Direct oral anticoagulant
F1+2: Prothrombin fragment 1+2
IQR: Interquartile range
SFMC: Soluble fibrin monomer complex

